# COVID-19 Vaccines for Children with Developmental Disabilities: Parent Survey of Willingness and Concerns

**DOI:** 10.1101/2021.12.23.21267953

**Authors:** Karen Bonuck, Suzannah Iadarola, Qi Gao, Joanne F. Siegel

**Affiliations:** Department of Family and Social Medicine, Division of Research, Department of Pediatrics, Division of Developmental Medicine, Einstein College of Medicine- Montefiore Medical Center, 1300 Morris Park Avenue, Bronx NY 10461; Department of Pediatrics, Division of Developmental and Behavioral Pediatrics, University of Rochester Medical Center; Department of Epidemiology and Population Health, Einstein College of Medicine-Montefiore Medical Center; Department of Pediatrics, Division of Developmental Medicine, Einstein College of Medicine- Montefiore Medical Center

## Abstract

**Objective:** While 1-in-6 US children has a developmental disability (DD), and such children are disproportionately affected by COVID-19, little is known about their vaccination status. We surveyed US parents of children with DDs to ascertain willingness and concerns regarding COVID-19 vaccines.

**Methods:** An online survey was distributed to national, statewide, and regional DD networks from June-September 2021. (Vaccines were authorized for adolescents in May 2021.) We report associations between vaccine willingness and concerns and: race/ethnicity, child age, in-person schooling, routine/flu vaccinations, and DD diagnoses. Willingness was categorized as Got /Will Get ASAP (high), Wait and See/Only if Required, or Definitely Not.

**Results:** 393 parents (51.2% white) responded. Willingness differed by age (p<.001). High willingness was reported for 75.3%, 48.9%, and 38.1% of children aged 12-17, 6-11 and 0-5 years-old, respectively. Willingness differed by Autism diagnosis (p<.001) and routine and flu vaccination status (p<.01). Predominant concerns included side effects (89%) and children with disabilities not being in trials (79%). Less common concerns were: COVID not serious enough in children to warrant vaccine (22%) and misinformation (e.g., microchips, 5G, DNA changes) (24%). Concerns about vaccine safety differed by age (p<.05) and were highest for young children. In age-stratified adjusted models, Autism was positively associated with high willingness for 6-11year-olds (OR= 2.66, 95% CI= 1.12-6.35).

**Conclusion:** Parents of children with DD are more willing for them to receive COVID-19 vaccines, compared to the general population. While few factors predicted willingness to vaccinate, addressing safety and developmental concerns regarding young children is warranted.

## INTRODUCTION

SARS CoV-2 (COVID-19) poses a heightened risk for individuals with Intellectual and/or Developmental Disabilities (DD). “DD” includes disorders that: begin in childhood; impact physical, social and/or emotional development, and; may affect multiple body parts or systems. They span conditions such as autism spectrum disorder, cerebral palsy, Down syndrome, hearing loss, and attention deficit hyperactivity disorder (ADHD). COVID-specific disparities in outcome vary within IDD; Intellectual Disability (ID) is associated with higher infection and case-fatality rates in adults.^1^.^2^ while DDs more generally increase the likelihood of hospitalization associated with COVID-19. For example, people with co-morbid autism and ID were 9.3 times more likely to be hospitalized than people with COVID-19 without those diagnoses, while those with learning disabilities and autism (alone) were 3.8 and 3.6 times, respectively, more likely to be hospitalized than people without those diagnoses.^3^ Further, adults with disabilities are more likely to want-- but less likely to receive-- a COVID-19 vaccine, than their non-disabled peers, in part due to inaccessible systems.^4^

Approximately 1 in 6 children in the US has a developmental disability.^5^ Children with DD are disproportionately affected by COVID due to medical conditions that increase risk of severe illness, difficulty practicing preventive behaviors (i.e., mask-wearing, social distancing) and limited access to accessible vaccine settings.^6^ Amidst the pandemic, children with DDs have experienced worsening anxiety, ADHD, and Autism symptoms,^7,8^ along with increased medication usage.^9^ Given that children comprise 22% of the US population,^10^ vaccine uptake in children is critical to protecting themselves, as well as the community at large.^11^ While general (i.e., non-COVID-19) childhood vaccination rates in the US have slowly increased in the past decade, current vaccination rates are impeded by COVID-19’s impact on health care access and an eco-system of generalized vaccine hesitancy.^12^ Pre-pandemic, children with ID were less likely to have completed recommended vaccinations,^13,14^ as were children with Autism (and their younger siblings).^14^ Furthermore, even compared with parents of children with other chronic health or developmental conditions, parents of children with Autism express more vaccine concerns.^15^

Pre-existing concern regarding approved vaccines also influence intention towards COVID-19 vaccines for children. Surveys from 2020, found that 60-70% of US parents expressed willingness for their child to receive a COVID-19 vaccine.^16,17^ However, in an early 2021 survey of US parents, just 40% were willing to. Willingness increased with child age, while unwillingness was linked to concerns about the vaccine being too new, side effects, and lasting health problems.^18^ Among parents with children aged <= 12 years, just 49% in a national sample would vaccinate their youngest child (median age= 4.7 years). Not having a child in school/daycare and living in the South or Midwest was associated with increased hesitance.^19^ In this same survey with New York City parents, 62% would vaccinate their youngest child.^20^ Following the May 2021 emergency use authorization (EAU) of a vaccine for 12-15 year-olds in the US, ^21^ the Kaiser Family Foundation’s (KFF) Vaccine Monitor found increasing uptake and acceptance of vaccines for children. In September 2021, 52% of 12-17 year-olds had received the vaccine/or would as soon as possible (ASAP) vs. 42% in both May and June of 2021. Still, parents of just 34% for 5-11 year-olds and 23% for children under 5 years old expressed willingness to vaccinate their child.^22^

Children become infected with, get sick from, and transmit COVID-19;^23^ their rates of infection exceeded those of adults for the first time in August 2021.^24^ Since the 2021-22 school year began, 1-in-4 children had to quarantine due to possible school-related exposures.^22^ School closures, undertaken to reduce COVID-19 spread, adversely affect children’s health and well-being.^25^ Furthermore, though children with DDs are disproportionately affected by COVID-19, relatively little is known about parents/caregivers willingness and concerns regarding COVID-19 vaccines for their children. Thus, we conducted an online survey of parents of children with DDs during Summer-Fall 2021 to identify: a) willingness to vaccinate by demographic (i.e., diagnosis) and related risk factors (i.e., routine vaccination status), b) vaccine-related concerns by child age, and; c) associations between risk factors and high willingness to vaccinate, stratified by child age.

## METHODS

### Survey Development

This survey was adapted from the authors prior study of COVID-19 vaccine perceptions and intentions among adults with IDD, their caregivers, and professionals who work with them.^26^ For this parent/caregiver survey, many items were drawn directly from the prior instrument, including those used in the KFF surveys.^27^ Additional items geared to parents/caregivers were informed by the literature, and “Engage & Educate,” the team’s statewide campaign to promote vaccines among persons with DDs in New York State. These items included: school/ child care placement, preferred location for vaccination, and child-specific concerns about COVID-19 vaccines (e.g., effects on child development, fertility; interaction with required childhood vaccines), Data were analyzed by three age groups: a) 12-17 years, i.e., adolescents, for whom there was EAUs at the time of the survey, b) 6-11 years, i.e., school-age, who were the next age group up for EAU review, and c) <= 5 years, i.e., young children. After review by stakeholders, including parent- and self-advocates, and disability service providers, the survey was translated into the following languages of the Engage & Educate project’s community partners: Bengali, Chinese, Korean, and Spanish.

This project received a determination of “Non-Research” from both the Institutional Review of the Albert Einstein College of Medicine and from the Research Subjects Review Board at the University of Rochester.

### Survey Distribution

The survey was built into Research Electronic Data Capture (REDCap), a user-friendly, HIPAA compliant data capture system. In addition to the above languages, the survey was translated into American Sign Language (ASL). For the ASL version, all instructions and survey items were video recorded in ASL and then attached to each individual question. An electronic link with a plain language description of the survey in all languages was posted via national, statewide, and regional networks and social media channels, from June 3 thru September 21, 2021. The three University Centers of Excellence in Developmental Disabilities (UCEDDs) in New York State also disseminated the survey to their networks which included local health departments; service providers, Special Olympics New York, school districts, a 150-member consortium of congregate care providers, etc. Organizations serving persons with IDD who are Hispanic/Latino, Chinese, Korean, and South Asian also distributed the survey. Within NYS, the survey was shared via listservs and social media pages specific to parents of children with disabilities (e.g., parent advisory groups, parent leadership cohorts, special education parent-teacher associations). The survey was also disseminated via a national disability network.

### Survey Items

#### Demographics and Placement Preferences

Individuals self-selected into the survey by indicating that they were a “caregiver who makes medical decisions for a child (under age 18) with a developmental disability,” and were asked to complete the survey regarding one such index child. Participants reported on basic demographic information about their children including age band (i.e., 12-17; 6-11; 5 or younger), diagnoses provided, child hearing status, race and ethnicity, whether the child is up to date for current vaccines (including flu vaccine), and state of residence. Both 1) *current* school/ childcare placement and 2) *preferred* school/ childcare placement modality *for fall* were assessed with the following options: “100% in person,” “100% remote,” “hybrid (a mix of in-person and remote),” “in my home (provided by someone else,” “in someone else’s home,” “center-based,” and “homeschooled.” Any setting outside of home was classified as ‘in-person.’

#### Vaccine Willingness

Willingness to receive the COVID-19 vaccine was assessed with the following KFF item (additional instructions from the current survey italicized): “*For your child aged [12-17; 6-11; 5 or younger]:* How likely would you be to get the COVID-19 vaccine if it were offered for free?” Response options included: “Already got (shown only for children aged 12-17),” “Will get as soon as possible,” “Wait and see,” “Only if required,” and “Definitely Not.” We combined the first two responses into a “Got/Get ASAP” (i.e., “highly willing”) category. The next two categories were combined into a “Waiting/If Required” (i.e., “not yet willing”) category. Respondents who fell within either the final “Do Not Intend” or “Waiting/If Required” category were shown an additional set of questions related to concerns about the vaccines.

#### Vaccine Concerns

Those who did not report high intention around the vaccine were asked to rate potential reasons for concern as a “big reason,” “little reason,” or “not a reason.” Concerns were grouped into overarching themes: general COVID concerns and specific child/family concerns about the vaccine, age-related concerns (for children aged 5 and younger only, including that the child is too young and the child is already getting too many vaccines at their age), and general concerns about vaccines.

#### Preferred Vaccine Location

Parents were asked to rank order where they would prefer their child to be vaccinated for COVID-19, including pediatrician or family doctor, public vaccination site, grocery store, child’s school, mobile vaccine unit, drive-through site, in-home service, and faith institution.

### Analysis

Descriptive analysis was performed on parent and child demographics and COVID-19 vaccine related items. Mean and standard deviation were presented for the continuous variables, while count and percentage were presented the categorical variables. Between group difference were tested by using student’s t-test or Chi-square test. To examine the association between willingness to get vaccine and potential risk factors, willingness was dichotomized into “Got/Get ASAP” and “Else”. A logistic regression model was used with “Got/Get ASAP” as the dependent variable, and race/ethnicity, school plan, flu vaccine status, comorbidities as independent variables, stratified by age groups. As actual and planned EAU status varied across the three age groups, we expected that risk factor effects would also vary by age, so multivariate logistic regression models were run, stratified by age group. All analyses were performed by using SAS 9.4 (SAS Institute Inc., Cary, NC, USA).

## RESULTS

### Demographics and Willingness to Vaccinate (Table 1)

This diverse sample of n= 393 included parents of Asian (18.4%), Black/African American (8.5%) and Hispanic/Latina(o) (14.9%) descent, 56.8% of whose index child was <12 years old. Most respondents were from New York State (89.6% [not shown]). Autism was the predominant diagnosis (60.6%), followed by ADHD/Learning (30.3%), and Developmental Delay (24.2%). Though New York state announced in May 2021 that its schools would open for in-person learning in the Fall, 25% of parents weren’t planning for their child to attend school in-person in September.^28^ Most children’s routine vaccinations were up to date (94.1%), but only 70.2% of age-eligible children had received a flu shot in the last year. The top ranked vaccination settings, which varied by age (p<.01) were doctor’s office, other and home.

Willingness to vaccinate differed by race/ethnicity (p<.001). White and Asian parents were highly willing to vaccinate their child (68.2% and 59.4%, respectively), while 31.3% of Black/African-American and 21.4% of Hispanic/Latino(a) parents did not intend to. Willingness also differed by child age (p<.001). High willingness was reported by parents of 75.3% of adolescents, 48.9% of school-aged children, and 38.1% of young children. Autism was the only diagnosis associated with willingness to vaccinate (p<.01). Parents of children with autism (vs. without Autism) were more than twice as likely to “definitely not” want their child vaccinated (18.0% vs. 8.4%). Routine and flu vaccination status were both associated with willingness to vaccinate (p<.01); school plans and preferred vaccination site were not.

### Concerns About Getting Vaccine (Table 2)

Among parents who weren’t highly willing to vaccinate their child (n= 165), the most frequently reported “Big Reasons” for concern were side effects (89%), non-inclusion of children with disabilities in clinical trials (78.8%), vaccine being too new and wanting to see effect on others (75.5%), effect on child’s development (67.7%) and not wanting child to be used as an experiment (64%). Concerns related to getting COVID from the vaccine (19.6%), the seriousness of COVID in children (22.1%), vaccines containing microchips/5G/changing DNA (23.9%) and needle anxiety (29.4%) were less common. Among parents of children aged 0-5 years, 75% indicated that their child was too young for the vaccine.

There were few significant age-related differences across the 16 listed concerns (18 concerns for 0-5 year olds). These included concerns about the vaccine’s safety (p<.05), which was highest for children aged 0-5 years (35.9%) compared with 6-11 (33.3%) or 12-17 (30.8%) year-olds. Lack of trust in vaccines generally (p<.001) and in the health care system (p<.001) was higher for the young children and adolescents, though sample sizes were small.

### Predictors of ‘Got/Will Get ASAP’ Vaccine (Table 3)

In adjusted models, compared to white parents, the odds of high willingness to vaccinate 0-5 year old children were lower for Asian (OR= 0.11; 95% CI: 0.02-0.81), Black (OR= 0.06; 96% CI:0.01-0.43) and Hispanic (OR= 0.11; 95% CI: 0.02-0.58) parents. No other variables were significant in the model for the youngest children. Among parents of 6-11 year-olds, *reduced* odds of being highly willing were reported by Hispanic (vs. white) parents (OR= 0.26; 95% CI: 0.07-0.99), while *increased* odds were reported by parents whose child has Autism (OR= 2.66; 95% CI: 1.12-6.35). There were no significant predictors in the model for 12-17 year olds.

## DISCUSSION

This paper reports on novel data from nearly 400 parents of children with DDs about COVID-19 vaccines. Compared to the general population, parents of children with DDs appear more willing for their children to be vaccinated. (For context, study data were collected *after* vaccine EAU for 12-15-year-olds in May 2021-- which expanded the December 2020 EAU for >= 16 year-olds-- and *before* the October 2021 EAU for 6-11-year-olds.^21^) Parents of 75% of 12-17 year-olds got or would get ASAP the vaccine for their child with a DD, versus 52% in the September 2021 KFF Vaccine Monitor. Similarly, willingness to get the vaccine ASAP was higher for school-age children in our study than in the KFF survey(49% vs. 34%), as well as in the youngest children (38% vs. 23%).^22^ Further, as of mid-December 2021, 19% of 5-11 year-olds and 62% of 12-17 year-olds in the US had received at least one vaccine dose.^29^ These proportions are lower than our “get/get ASAP” study data from months earlier at these ages, high vaccine demand for children with DD.

To meet this demand, the top ranked vaccination settings were doctor’s offices, followed by public sites and schools. This ordering parallels New York State’s roll-out, which prioritized doctor’s offices and community pharmacies. Unlike health care settings that care for children with IDD, not all community sites and schools have IDD trained staff and behavioral supports needed to provide positive vaccine experience for children with IDD (e.g., visual schedules, social stories, and sensory distraction materials). Efforts are also needed to increase access to healthcare visits (e.g., flexible scheduling, transportation assistance) and to emphasize that COVID-19 vaccinations not be devalued until their child’s next well-child visit. Additionally, vaccine outreach and education must be tailored to the DD population’s concerns about side effects, non-inclusion of children with disabilities in vaccine trials, and effects on their development. Notably, these DD-specific concerns overlap with those identified for adults with DD, such as representation of persons with DD in vaccine trials and not wanting to be used as not wanting to be “used as experiment.”^30^ These findings underscore broader issues about the lack of inclusion of persons with DDs in disease surveillance and research. Finally, outreach to families of color is needed to address perceptions of a health care system that has often been difficult to access, not met their needs, and oftentimes failed to deliver respectful care.

Fewer children with autism and other DDs are current with their recommended vaccinations,^13-15^ largely due to long debunked myths.^15^ Not surprisingly, study parents not intending to vaccinate their child (albeit just 14% of the sample), were three times as likely to report their child was not up to date with routine vaccinations or the flu shot. Findings by specific diagnosis were nuanced. Unadjusted for age or other factors, *high* willingness was comparable for children with autism (59%), sensory/other (61%) and ADHD/Learning (62%) diagnoses, while *low* willingness (“do not intend”) was higher for parents of children with (vs. without) autism: 18% vs. 8%. This finding suggests that parents of children with autism may have less hesitance towards COVID-19 versus other vaccines. Indeed, in age-adjusted multivariate models, autism was associated with *higher* odds of school-age children getting the vaccine, but this was the *only* significant finding for diagnosis in age-stratified models. Thus, parents’ decisions about COVID-19 vaccines for their children with DDs are multi-factorial and may not mirror prior data about vaccines and autism.

In terms of race/ethnicity, a higher *overall* percentage of Black and Hispanic parents were not willing to vaccinate their child. Similarly, some people of color in the US were initially more vaccine hesitant—though gaps with white non-Hispanics have largely narrowed.^29^ There is valid and historical mistrust in the health care system among communities of color in this country, which likely affected initial hesitance. In response, culturally and linguistically competent outreach and education sought to target mistrust, and likely reduced hesitance. In contrast to our “Table 1 findings,” in age-stratified models there were no significant differences in “high willingness” by any race/ethnicity for adolescents, and only lower odds for Hispanic (vs. white) for school-age children, In contrast, white parents of young children were significantly more likely to get the vaccine ASAP than all other race/ethnic groups. These finding suggest that parents of color are balancing health care system mistrust and concerns about the vaccine, with short- and long-term concerns about their child’s health, and the COVID-19 vaccine’s potential for families to get back to or maintain their routines.

**Table 1.**
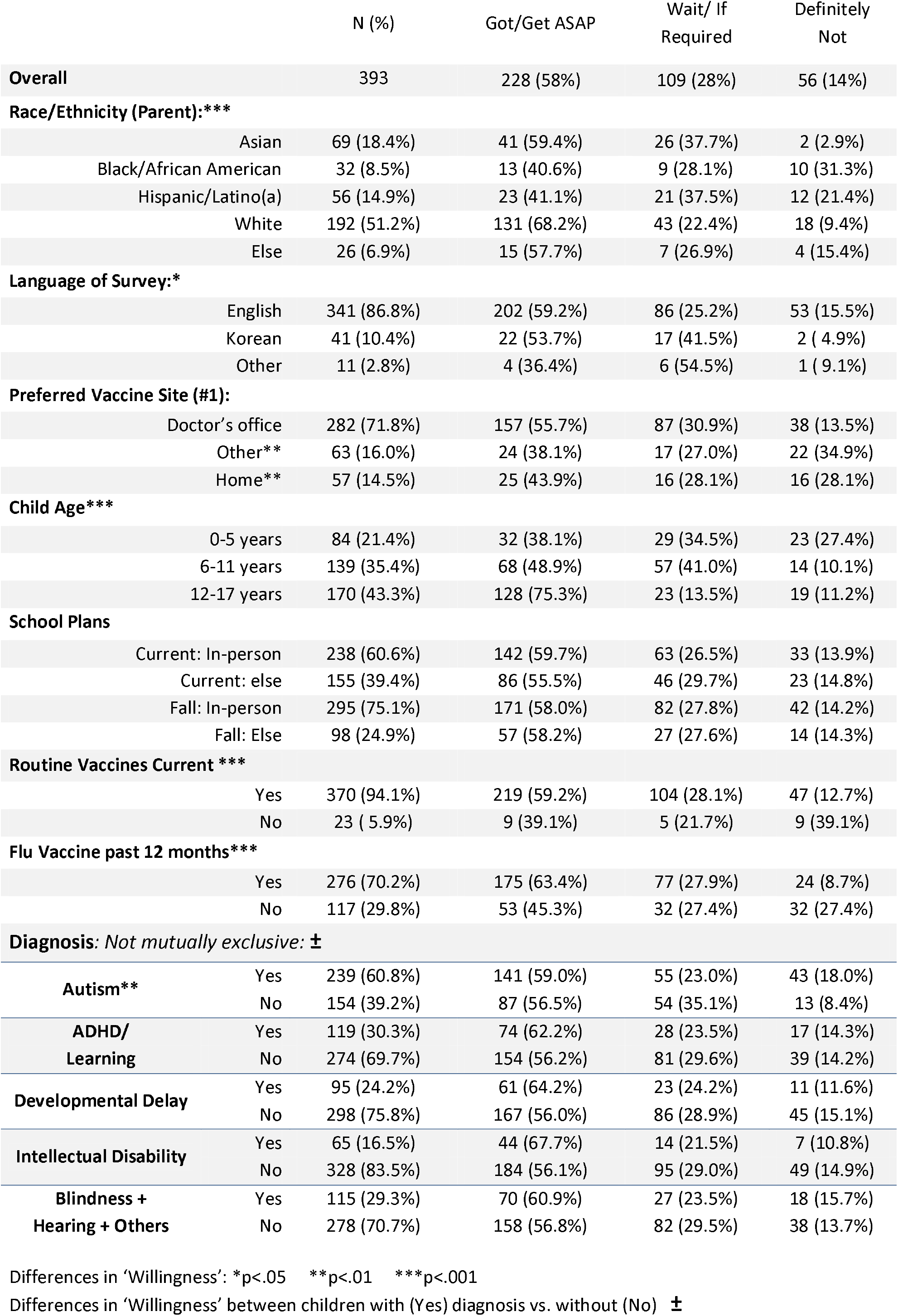
Demographics and Willingness to Vaccinate.

**Table 2:** Concerns About Getting Vaccine: Total Sample and by Age.

| Overall | N (%) | 0-5 | 6-11 | 12-17 |
| --- | --- | --- | --- | --- |
|  | N=165 | N=52 | N=71 | N=42 |
| **COVID Concerns:** |  |  |  |  |
| *Side Effects* | 146 (89.0%) | 48 (32.9%) | 63 (43.2%) | 35 (24.0%) |
| *COVID vaccine not safe\** | 78 (47.6%) | 28 (35.9%) | 26 (33.3%) | 24 (30.8%) |
| *Children w disabilities not in trials* | 130 (78.8%) | 46 (35.4%) | 53 (40.8%) | 31 (23.8%) |
| *Too new, want to see effect on others* | 123 (75.5%) | 40 (32.5%) | 53 (43.1%) | 30 (24.4%) |
| *COVID not serious enough in children to need vaccine* | 36 (22.1%) | 10 (27.8%) | 14 (38.9%) | 12 (33.3%) |
| *Vaccine might contain microchip, 5G, change DNA* | 39 (23.9%) | 15 (38.5%) | 12 (30.8%) | 12 (30.8%) |
| *Won't decide until trials in this age group* | 73 (59.8%) | 32 (43.8%) | 41 (56.2%) | N/A |
| **Vaccine Concerns:** |  |  |  |  |
| *Don't trust vaccines in general\*\** | 37 (22.6%) | 16 (43.2%) | 6 (16.2%) | 15 (40.5%) |
| *Don't trust health care system\*\** | 32 (19.8%) | 11 (34.4%) | 6 (18.8%) | 15 (46.9%) |
| **Specific Child/Family Concerns:** |  |  |  |  |
| *Effects on child's development* | 111 (67.7%) | 39 (35.1%) | 47 (42.3%) | 25 (22.5%) |
| *Effects on fertility* | 69 (42.3%) | 22 (31.9%) | 33 (47.8%) | 14 (20.3%) |
| *Child might get COVID from vaccine* | 32 (19.6%) | 10 (31.3%) | 14 (43.8%) | 8 (25.0%) |
| *Don't want child used as 'experiment'* | 105 (64.0%) | 39 (37.1%) | 40 (38.1%) | 26 (24.8%) |
| *Religious or cultural reasons* | 19 (11.7%) | 7 (36.8%) | 5 (26.3%) | 7 (36.8%) |
| *Needle anxiety* | 48 (29.4%) | 18 (37.5%) | 18 (37.5%) | 12 (25.0%) |
| *Vaccine site won't support child's needs* | 53 (32.3%) | 19 (35.8%) | 23 (43.4%) | 11 (20.8%) |
| *My child is too young for this vaccine* | 39 (75.0%) | 39 (100%) | N/A | N/A |
| *Child is getting too many vaccines now* | 15 (30.0%) | 15 (100%) | N/A | N/A |

Age differences: \*p<.05 \*\*p<.01

**Table 3:** Predictors of “Got/Get ASAP” Vaccine for Child.

| Level | 0-5 Years |  |  | 6-11 years |  |  | 12-17 years |  |  |
| --- | --- | --- | --- | --- | --- | --- | --- | --- | --- |
|  | Odds Ratio | Lower 95% CI | Upper 95% CI | Odds Ratio | Lower 95% CI | Upper 95% CI | Odds Ratio | Lower 95% CI | Upper 95% CI |
| **Race (vs white)** |  |  |  |  |  |  |  |  |  |
| Asian | 0.11* | 0.02 | 0.81 | 0.92 | 0.31 | 2.73 | 1.18 | 0.30 | 4.61 |
| Black | 0.06** | 0.01 | 0.43 | 1.06 | 0.20 | 5.48 | 0.79 | 0.13 | 4.72 |
| Hispanic | 0.11** | 0.02 | 0.58 | 0.26* | 0.07 | 0.99 | 0.49 | 0.16 | 1.55 |
| Others | 0.08* | 0.01 | 0.89 | 1.31 | 0.31 | 5.60 | 1.04 | 0.22 | 5.01 |
| **Current Schooling:** |  |  |  |  |  |  |  |  |  |
| Any In-person (vs. None) | 1.11 | 0.31 | 3.95 | 1.82 | 0.69 | 4.82 | Table | 0.35 | 2.29 |
| **Fall School Plan:** |  |  |  |  |  |  |  |  |  |
| Any In-person (vs. None) | 0.91 | 0.19 | 4.35 | 0.70 | 0.25 | 1.97 | 0.92 | 0.31 | 2.69 |
| **Flu Vaccine, Recent:** |  |  |  |  |  |  |  |  |  |
| Got Vaccine (vs. Not) | 2.68 | 0.50 | 14.45 | 2.32 | 0.93 | 5.79 | 5.57 | 2.39 | 12.96 |
| **Diagnosis, (vs. No)** |  |  |  |  |  |  |  |  |  |
| Autism | 2.30 | 0.56 | 9.43 | 2.66** | 1.12 | 6.35 | 1.14 | 0.44 | 2.94 |
| ADHD | 0.11 | 0.01 | 1.27 | 1.37 | 0.63 | 2.97 | 0.67 | 0.27 | 1.71 |
| Developmental Delay | 2.04 | 0.53 | 7.79 | 0.95 | 0.37 | 2.45 | 1.64 | 0.49 | 5.50 |
| Intellectual Disability | 2.13 | 0.23 | 19.91 | 1.51 | 0.51 | 4.51 | 0.99 | 0.30 | 3.26 |
| Vision/Hearing/Other | 0.57 | 0.11 | 2.86 | 1.95 | 0.72 | 5.26 | 0.62 | 0.23 | 1.71 |

\*p<.05 \*\*p<.01

To our knowledge, this is the first such parent/caregiver data in the DD population, thus filling a gap in knowledge for this vulnerable population. We note that national data track COVID-19 vaccination rates only for adults (i.e., >= 18 years) with disabilities, and do not track rates for people with DDs.^29^ Another strength is the sample’s race/ethnic and linguistic diversity, including nearly half persons of color and 13% who completed the survey in a language other than English. In addition, inclusion of vetted items, such as the KFF Vaccine Monitor classifications for willingness to vaccinate, enables comparisons. A main limitation is the convenience sample, which may bias towards families connected to the DD networks from which we recruited. Relatedly, most respondents were from New York State. Thus, findings may not be generalizable to the US given pre-pandemic demographic differences, and state-specific COVID-19 infection and morbidity rates, and guidelines. Notably, 49% of US parents in a prior survey intended to vaccinate their youngest child^19^ compared with 62% of New York City parents using the same survey.^20^ Finally, the sample may exclude parents who were unable to access and navigate a web-linked survey.

### Conclusion

Children with DDs are vulnerable to both the health risks posed by COVID-19, as well as the pandemic’s impact on developmental services, in-person schooling, and socialization opportunities. The relatively high willingness of parents of children with DDs to vaccinate their children is encouraging, though trust will need to be engendered among parents of young children of color to actualize these intentions. Additional monitoring data are needed to track COVID-19 disease, vaccine intentions and vaccination rates in children.

## Data Availability

All data produced in the present study are available upon reasonable request to the authors

## Acknowledgments

The authors would like to thank all of our language and ASL community partners for assisting with survey distribution.

